# Implementation of the Short-Form 36 (SF-36) in childhood cancer survivors: An analysis on measurement properties across 5 European countries

**DOI:** 10.1101/2024.01.23.24301414

**Authors:** Katja Baust, Gabriele Calaminus, Claire Berger, Julianne Byrne, Desiree Grabow, Marry M. van den Heuvel-Eibrink, Melanie Kaiser, Tomas Kepak, Leontien C.M. Kremer, Jarmila Kruseova, Rahel Kuonen, Katharina Roser, Claudia Spix, Heleen Maurice-Stam, Claudia E. Kuehni, Grit Sommer

## Abstract

**Purpose:** The Short Form-36 (SF-36) is widely used in many research contexts and cultures to assess health-related quality of life (HRQOL). We investigated the measurement properties of the SF-36 in a large cohort study among childhood cancer survivors living in 5 European countries.

**Methods:** The PanCareLIFE project includes adult survivors of childhood cancer living in the Czech Republic, France, Germany, The Netherlands, and Switzerland. We invited 19,268 survivors aged >18 years, and 10,077 (53%) returned questionnaires. Of these, 9,871 (98%) were included in the analyses. We assessed HRQOL with the SF-36 version 1 (V1) or version 2 (V2) and investigated its performance. We analysed data completeness, floor and ceiling effects, item-internal consistency, item-discriminant validity, reliability, and scaling assumptions focusing on country-and version-specific differences.

**Results:** Data completeness was high but differed between countries (90% in the Czech Republic to 96% in France). Floor effects were negligible, but ceiling effects in 5 out of 8 SF-36 scales were present (50-70%). V2 had slightly fewer ceiling effects than V1 in 2 scales. Item-internal consistency, item-discriminant validity, and reliability were good. Scaling assumptions were met, with 4 scales representing strong mental health content, and the other 4 representing strong physical health content.

**Conclusion:** The SF-36 (V1 and V2) provided robust measurement properties among childhood cancer survivors, and differences in study design across the 5 European countries did not affect measurement properties. Researchers need to take into account that ceiling effects exist which might limit sensitivity among participants who fared particularly well.

**PLAIN ENGLISH SUMMARY:** This study investigated how well the Short Form-36 (SF-36), a commonly used tool for assessing health-related quality of life, performs in childhood cancer survivors from five different European countries. We looked at the amount of survivors answering to all of the SF-36 questions, how many indicated the most positive or the most negative answer category, whether the questions were consistent within the different aspects of the questionnaire and how consistent and dependable it measured quality of life in the survivors. We found that overall, the SF-36 measured health-related quality of life well in all the countries. This is reassuring for researchers considering its use in studies involving multiple centers across Europe.

## MAIN TEXT

## INTRODUCTION

Late effects after childhood cancer and its treatment can affect long-term health-related quality of life (HRQOL) [1–10]. The Short-Form 36 (SF-36) is a widely used instrument to assess HRQOL [11–13]. The childhood cancer survivor studies in the United States, the United Kingdom, Switzerland, and The Netherlands implemented the SF-36 as a long-term outcome measure [3; 6; 7; 9]. Most studies used version 1 (V1) of the SF-36 [13] and a few studies used version 2 (V2) [11] since its introduction in the late 1990s. Versions differ mainly by the number of response categories and wordings of several questions [11].

Since SF-36 measurement properties vary by version and cultural background of the population, researchers should evaluate the structural validity for a specific study population before interpreting questionnaire results [14; 15]. Both V1 and V2 of the SF-36 showed good structural validity among general population samples and a broad range of cohorts with chronic diseases, including multiple sclerosis, rheumatoid conditions, and HIV [15–24]. Among more than 10,000 adult survivors from the British Childhood Cancer Survivor Study (BCCSS), the SF-36 V1 provided overall valid and reliable psychometric criteria [25]. Such reassuring results prompted the European Seventh Framework project PanCareLIFE collaborators to use the SF-36 (either V1 or V2) for assessing HRQOL in a pooled sample of adult survivors of childhood cancer from 5 European countries: Czech Republic, France, Germany, The Netherlands, and Switzerland [26]. In this study, we evaluated measurement properties of the SF-36 for a cohort of nearly 10,000 survivors of childhood cancer in Europe, since cultural differences between countries and different SF-36 versions possibly affect HRQOL result interpretation. We aimed to compare differences between countries, including data completeness, floor and ceiling effects, item-internal consistency, item-discriminant validity, reliability, and scaling assumptions focusing on country-and version-specific differences.

## METHODS

### Study and study population

We used data from the European PanCareLIFE framework (grant agreement no. 602030). Five European countries (Czech Republic, France, Germany, The Netherlands, and Switzerland) contributed HRQOL and clinical data from adult survivors of childhood cancer. Eligible participants were 1) younger than age 18 at the time of diagnosis; 2) residents of the respective participating country at the time of diagnosis; 3) diagnosed with cancer according to the International Classification of Childhood Cancer, 3rd edition (ICCC-3) [27] or Langerhans cell histiocytosis; 4) who were alive at least 5 years after cancer diagnosis; 5) age 18 or older when they completed questionnaires; and 6) not undergoing treatment for cancer at time of study [26]. All survivors participated in regional or national population–based cohort studies. In the Czech Republic, data came from the university hospitals in Prague and Brno, which cover childhood cancer survivors in 95% of the country and in France from the regional population–based registry of the Rhone-Alps region. Germany, The Netherlands, and Switzerland collected data through their own population– and hospital–based cohort studies with >95% national coverage: the German VIVE cohort [8], the Dutch Childhood Cancer Survivor Study (DCCSS LATER, previously named DCOG LATER) [28], and the Swiss Childhood Cancer Survivor Study (SCCSS) [29]. Study design, patient characteristics, and sampling methods for the overall cohort and for each country were published elsewhere [26].

In brief, participants completed the questionnaires between 2005–2017. Eligible childhood cancer survivors received questionnaires including the SF-36 and sociodemographic questions as part of their scheduled follow-up consultations (Czech Republic) or by postal mail in other participating countries. SF-36 questions were asked either at beginning of longer questionnaires (France, The Netherlands, and Switzerland) or at the end (Czech Republic and Germany). The Czech Republic, Germany, and The Netherlands used V1 of the SF-36; France and Switzerland used V2. Survivors who did not return completed questionnaires were reminded at least once. Clinical baseline data came from respective cancer registries or hospital databases of each country. **Supplemental table 1** describes participant characteristics stratified by SF-36 version. Country-specific characteristics were published elsewhere [26]. All data contributed to PanCareLIFE were pseudonymized. All survivors provided either written informed consent or implied consent or assent by returning the questionnaire.

### Short Form-36 (SF-36)

The SF-36 includes 35 items covering 8 scales: physical functioning (PF, 10 items), role-limitations due to physical problems (role physical, RP, 4 items), bodily pain (BP, 2 items), general health (GH, 5 items), vitality (VT, 4 items), social functioning (SF, 2 items), role-limitations due to emotional problems (role emotional, RE, 3 items) and mental health (MH, 5 items). One additional SF-36 item describes a subjective change in health over a one year time period, but it is not part of this study. From the 8 scales, 2 summary scores—physical component summary (PCS) and mental component summary (MCS)— are derived by factor analysis. Items use a Likert scale with a maximum range of possible answers from 1 to 6. The 2 SF-36 versions differ in layout, font size and bolding, wording, and number of response choices. Wording changes from V1 to V2 include shortening and simplifying instructions and item text. Response scale changes refer to replacing dichotomous (RP and RE) and some 6-level response choices (MH and VT) with 5-level response choices [11].

According to the User’s Manual of the SF-36 V2, we translated answers into percentage scores ranging from 0–100 [11]. Higher scores indicate better HRQOL. We transformed all scales from V2 to V1 in line with the User’s Manual [11]. Since normative data were not available for all participating countries, we used normative data from a representative population in Germany from 1998 [30]. This is a well-described sample including the largest number of participants from a country participating in our study and it used V1, which is also mostly used in our sample. It also allowed comparing measurement properties between countries. We converted raw scores into T-scores (mean=50, SD=10), according to the German normative data stratified for age and gender from 1998 [30].

### Assessed criteria

Terwee et al. proposed quality criteria for evaluating health status questionnaires [31]. Based on these recommendations, we evaluated a) data completeness; b) floor and ceiling effects; c) item-internal consistency; d) item-discriminant validity; e) reliability; and f) checked scaling assumptions of the 2 SF-36 summary scores, MCS and PCS. We stratified analyses for data completeness by country, for all other criteria by version, country, gender, cancer diagnosis and age at survey, and for criterion f) by version of the SF-36.

### Statistical analyses

#### Data completeness

We described missing data by tabulating frequencies of survivors with unanswered single items on the SF-36 and calculating percentages of missing data for each item.

#### Floor and ceiling effects

We measured the extent responses reached the upper and lower limit of the measurement range by calculating percentages of survivors with the lowest and highest scores for each scale. Floor and ceiling effects were present if >15% of participants responded with the lowest or highest score, respectively [31].

#### Item-internal consistency

We assessed whether items measured similar constructs as presumed by the developers of the SF-36. Within each of the 8 SF-36 scales, we calculated correlations between each item and its respective related scale (item-rest correlations). We considered correlations >0.4 satisfactory for item-internal consistency [13].

#### Item-discriminant validity

To determine item-discriminant validity, we checked whether the correlation between an item and its hypothesized scale was higher than the correlations between that item and all other scales [32]. We subtracted correlations between each item and their unrelated scales (item-scale correlations) from the item-rest correlations of the same item calculated. We considered differences of >2 standard errors between item-rest correlations and item-scale correlations as scaling success, indicating the item correlated higher with its hypothesized scale than with the other—supposedly unrelated—scales. We calculated success rates as the proportion of comparisons with scaling success divided by the total number of comparisons [13].

#### Scale reliability

We calculated Cronbach’s alpha for all scales including the respective items of the scale. Terwee et al. proposed a Cronbach’s alpha coefficient between 0.7 and 0.95 as satisfactory for a good internal consistency [31], and the User’s Manual of the SF-36 V2 a threshold of ≥0.7 for acceptable reliability [11].

#### Scaling assumptions of the summary scores

We performed principal component analyses and subsequent orthogonal varimax rotation and calculated factor loadings—correlations of each scale with the PCS and MCS—and estimated communality and uniqueness for V1 and V2 [23]. Communality is the variance of each scale explained by the PCS and MCS; uniqueness is the unexplained variance. We considered correlations of r>0.7 as strong and of r<0.3 as weak [13]. We expected strong correlations of the physical health scales PF, RP, and BP with the PCS and weak correlations with the MCS and strong correlations of the mental health scales MH and RE with the MCS and weak correlations with the PCS. We expected GH, VT, and SF scales with values in between.

## RESULTS

### Data completeness

Out of 24,993 eligible survivors, we contacted 19,268 survivors and 10,077 (53%) returned questionnaires (**Table 1**). Details on the eligible survivors and on those who were contacted for the study were published elsewhere [26]. In brief, the largest cohort of eligible survivors was in Germany (n=12,144); the smallest in France (n=1,060). Participation ranged from 65% (1,029/1,576) in the Czech Republic to 45% (385 out of 851 of contacted survivors) in France. Of the 10,077 survivors who returned questionnaires, 9,300 (92%) answered all items of the SF-36 and 9,871 (98%) answered at least half of the SF-36 items within a scale, which sufficed for scoring the SF-36 [13]. We found differences in completeness between countries (**Table 1**). France had the highest percentage of survivors who completed all items (377/391, 96%) and the Czech Republic had the lowest (979/1,085, 90%). In Switzerland, 0.3% (5/1,590) of participants answered less than half of the questions in at least 1 scale; in the Czech Republic, the figure was highest with 5% (56/1085). Data completeness was generally high, and the percentage of missing data never exceeded 4% for a single item. Items 8 (BP), 11b and 11d (GH), and 9a (VT) had the most missing values, while items 3h and 3i (PF) had the fewest missing values. We did not observe specific patterns in completeness for the two different versions of the SF-36.

**Table 1:** Missing data of SF-36 items for all survivors who answered at least 1 item of the SF-36 for all countries ned and stratified by country.

|  | All countries<br>combined<br>N=10,077 |  | Czech<br>Republic<br>N=1085 |  | France<br>N=391 |  | Germany<br>N=4835 |  | The Nether-<br>lands<br>N=2176 |  | Switzerland<br>N=1590 |  |
| --- | --- | --- | --- | --- | --- | --- | --- | --- | --- | --- | --- | --- |
|  | n | % | n | % | n | % | n | % | n | % | n | % |
| Physical functioning (PF) |  |  |  |  |  |  |  |  |  |  |  |  |
| Item 3a | 83 | 0.8 | 37 | 3.4 | 4 | 1 | 29 | 0.6 | 11 | 0.5 | 2 | 0.1 |
| Item 3b | 68 | 0.7 | 32 | 3 | 3 | 0.8 | 22 | 0.5 | 8 | 0.4 | 3 | 0.2 |
| Item 3c | 74 | 0.7 | 36 | 3.3 | 3 | 0.8 | 25 | 0.5 | 7 | 0.3 | 3 | 0.2 |
| Item 3d | 67 | 0.7 | 30 | 2.8 | 3 | 0.8 | 23 | 0.5 | 7 | 0.3 | 4 | 0.3 |
| Item 3e | 66 | 0.7 | 32 | 3 | 3 | 0.8 | 23 | 0.5 | 7 | 0.3 | 1 | 0.1 |
| Item 3f | 70 | 0.7 | 32 | 3 | 3 | 0.8 | 25 | 0.5 | 9 | 0.4 | 1 | 0.1 |
| Item 3g | 67 | 0.7 | 32 | 3 | 3 | 0.8 | 23 | 0.5 | 8 | 0.4 | 1 | 0.1 |
| Item 3h | 63 | 0.6 | 31 | 2.9 | 3 | 0.8 | 23 | 0.5 | 6 | 0.3 | 0 | 0 |
| Item 3i | 63 | 0.6 | 31 | 2.9 | 3 | 0.8 | 22 | 0.5 | 6 | 0.3 | 1 | 0.1 |
| Item 3j | 67 | 0.7 | 33 | 3 | 4 | 1 | 22 | 0.5 | 8 | 0.4 | 0 | 0 |
| Role physical (RP) |  |  |  |  |  |  |  |  |  |  |  |  |
| Item 4a | 83 | 0.8 | 29 | 2.7 | 7 | 1.8 | 40 | 0.8 | 7 | 0.3 | 0 | 0 |
| Item 4b | 83 | 0.8 | 29 | 2.7 | 5 | 1.3 | 39 | 0.8 | 10 | 0.5 | 0 | 0 |
| Item 4c | 93 | 0.9 | 27 | 2.5 | 5 | 1.3 | 45 | 0.9 | 11 | 0.5 | 5 | 0.3 |
| Item 4d | 97 | 1 | 27 | 2.5 | 6 | 1.5 | 46 | 1 | 13 | 0.6 | 5 | 0.3 |
| Bodily pain (BP) |  |  |  |  |  |  |  |  |  |  |  |  |
| Item 7 | 111 | 1.1 | 27 | 2.5 | 5 | 1.3 | 73 | 1.5 | 5 | 0.2 | 1 | 0.1 |
| Item 8 | 133 | 1.3 | 27 | 2.5 | 7 | 1.8 | 83 | 1.7 | 12 | 0.6 | 4 | 0.3 |
| General health (GH) |  |  |  |  |  |  |  |  |  |  |  |  |
| Item 1 | 66 | 0.7 | 18 | 1.7 | 2 | 0.5 | 43 | 0.9 | 2 | 0.1 | 1 | 0.1 |
| Item 11a | 113 | 1.1 | 28 | 2.6 | 3 | 0.8 | 57 | 1.2 | 20 | 0.9 | 5 | 0.3 |
| Item 11b | 126 | 1.3 | 30 | 2.8 | 3 | 0.8 | 67 | 1.4 | 21 | 1 | 5 | 0.3 |
| Item 11c | 100 | 1 | 27 | 2.5 | 3 | 0.8 | 52 | 1.1 | 14 | 0.6 | 4 | 0.3 |
| Item 11d | 133 | 1.3 | 30 | 2.8 | 3 | 0.8 | 72 | 1.5 | 16 | 0.7 | 12 | 0.8 |
| Vitality (VT) |  |  |  |  |  |  |  |  |  |  |  |  |
| Item 9a | 128 | 1.3 | 28 | 2.6 | 5 | 1.3 | 66 | 1.4 | 20 | 0.9 | 9 | 0.6 |
| Item 9e | 114 | 1.1 | 30 | 2.8 | 4 | 1 | 65 | 1.3 | 13 | 0.6 | 2 | 0.1 |
| Item 9g | 109 | 1.1 | 31 | 2.9 | 4 | 1 | 61 | 1.3 | 11 | 0.5 | 2 | 0.1 |
| Item 9i | 104 | 1 | 33 | 3 | 4 | 1 | 55 | 1.1 | 8 | 0.4 | 4 | 0.3 |
| Social functioning (SF) |  |  |  |  |  |  |  |  |  |  |  |  |
| Item 6 | 86 | 0.9 | 27 | 2.5 | 4 | 1 | 46 | 1 | 7 | 0.3 | 2 | 0.1 |
| Item 10 | 110 | 1.1 | 29 | 2.7 | 5 | 1.3 | 64 | 1.3 | 8 | 0.4 | 4 | 0.3 |
| Role emotional (RE) |  |  |  |  |  |  |  |  |  |  |  |  |
| Item 5a | 90 | 0.9 | 27 | 2.5 | 7 | 1.8 | 47 | 1 | 8 | 0.4 | 1 | 0.1 |
| Item 5b | 88 | 0.9 | 25 | 2.3 | 6 | 1.5 | 46 | 1 | 9 | 0.4 | 2 | 0.1 |
| Item 5c | 101 | 1 | 26 | 2.4 | 6 | 1.5 | 54 | 1.1 | 14 | 0.6 | 1 | 0.1 |
| Mental health (MH) |  |  |  |  |  |  |  |  |  |  |  |  |
| Item 9b | 99 | 1 | 29 | 2.7 | 4 | 1 | 54 | 1.1 | 3 | 0.4 | 3 | 0.2 |
| Item 9c | 102 | 1 | 29 | 2.7 | 4 | 1 | 53 | 1.1 | 14 | 0.6 | 2 | 0.1 |
| Item 9d | 100 | 1 | 31 | 2.9 | 4 | 1 | 48 | 1 | 13 | 0.6 | 4 | 0.3 |
| Item 9f | 98 | 1 | 28 | 2.6 | 4 | 1 | 50 | 1 | 13 | 0.6 | 3 | 0.2 |
| Item 9h | 109 | 1.1 | 31 | 2.9 | 4 | 1 | 62 | 1.3 | 9 | 0.4 | 3 | 0.2 |
| Incomplete in ≥1<br>item | 777 | 7.7 | 106 | 9.8 | 14 | 3.6 | 469 | 9.7 | 114 | 5.2 | 74 | 4.7 |
| All items complete | 9300 | 92.3 | 979 | 90.2 | 377 | 96.4 | 4366 | 90.3 | 2062 | 94.8 | 1516 | 95.4 |
| ≥50% missings in<br>≥1 scale | 206 | 2 | 56 | 5.2 | 6 | 1.5 | 109 | 2.3 | 30 | 1.4 | 5 | 0.3 |
| <50% missings per<br>scale | 9871 | 98 | 1029 | 94.8 | 385 | 98.5 | 4726 | 97.8 | 2164 | 98.6 | 1585 | 99.7 |

Abbreviations: BP: bodily pain, GH: general health, MH: mental health, PF: physical functioning, RE: role emotional, RP: role physical, SF: social functioning, SF-36, Short Form-36.

### Ceiling and floor effects

Ceiling effects were present in the 5 scales of PF, RP, BP, SF, and RE, yet not in the 3 scales of GH, VT and MH (**Table 2**). Among all scales, RP and RE had the highest ceiling effects (71% and 70%) (**Table 2**). Ceiling effects differed by version, country, gender, age at survey, and diagnostic group of the participants. Ceiling effects were higher in V1 (RP, 76%; RE, 74%) compared with V2 (RP, 63%; RE, 58%). Of all countries, Switzerland (PF, 71%; BP, 66%; SF, 59%) and Germany (RE and RP, 75%) had the highest ceiling effects. Among men ceiling effects were higher when compared with women (range 58%-77% vs. 55%-71%) (**Table 2**). Among all cancer diagnostic group, ceiling effects were high in the 5 scales of PF, RP, BP, SF, and RE, but less pronounced in survivors of CNS, hepatic and bone tumours. Ceiling effects were particularly high for young participants aged 25–35 years at survey and lower among older adults. Floor effects were rare **(Supplemental Table 2)**. We found them only in the scale of RP for bone tumour survivors and for survivors older than 40 years.

**Table 2:** Ceiling effects: Proportions of survivors with the highest possible score by country, SF-36 version, gender, cancer diagnosis, and age at survey.

|  | n | PF(%) | RP(%)* |  | BP(%) | GH(%) | VT(%)* |  | SF(%) | RE(%)* |  | MH(%)* |  |
| --- | --- | --- | --- | --- | --- | --- | --- | --- | --- | --- | --- | --- | --- |
|  |  |  | V1 | V2 |  |  | V1 | V2 |  | V1 | V2 | V1 | V2 |
| Overall cohort | 9871 | 51.26 | 72.54 | 62.54 | 56.28 | 8.72 | 2.44 | 3.35 | 53.01 | 73.52 | 58.12 | 3.72 | 4.57 |
| Country |  |  |  |  |  |  |  |  |  |  |  |  |  |
| Czech Republic | 1029 | 45.38 | 70.36 | n/a | 55.30 | 5.64 | 1.36 | n/a | 47.81 | 69.10 | n/a | 2.04 | n/a |
| France | 385 | 51.69 | n/a | 55.06 | 40.78 | 5.45 | n/a | 1.82 | 36.62 | n/a | 48.31 | n/a | 2.08 |
| Germany | 4726 | 52.31 | 75.43 | n/a | 60.85 | 8.72 | 2.20 | n/a | 56.79 | 74.63 | n/a | 3.24 | n/a |
| The Netherlands | 2146 | 44.69 | 67.19 | n/a | 41.94 | 6.52 | 3.49 | n/a | 45.90 | 73.21 | n/a | 5.59 | n/a |
| Switzerland | 1585 | 60.76 | n/a | 64.35 | 66.44 | 14.51 | n/a | 3.72 | 58.74 | n/a | 60.50 | n/a | 5.17 |
| Version |  |  |  |  |  |  |  |  |  |  |  |  |  |
| 1 | 7901 | 49.34 | 72.54 | n/a | 54.99 | 7.72 | 2.44 | n/a | 52.66 | 73.52 | n/a | 3.72 | n/a |
| 2 | 1970 | 58.98 | n/a | 62.54 | 61.42 | 12.74 | n/a | 3.35 | 54.42 | n/a | 58.12 | n/a | 4.57 |
| Gender |  |  |  |  |  |  |  |  |  |  |  |  |  |
| Male | 4725 | 58.48 | 76.30 | 65.84 | 61.38 | 10.60 | 3.51 | 4.74 | 57.86 | 76.50 | 65.59 | 4.58 | 5.65 |
| Female | 5146 | 44.64 | 69.16 | 59.14 | 51.99 | 7.00 | 1.49 | 1.94 | 48.56 | 70.53 | 50.56 | 2.95 | 3.47 |
| Cancer diagnosis (ICCC-3) |  |  |  |  |  |  |  |  |  |  |  |  |  |
| I Leukaemias | 3157 | 58.12 | 75.01 | 71.54 | 59.71 | 8.93 | 2.81 | 3.86 | 56.16 | 73.43 | 63.62 | 4.05 | 4.07 |
| II Lymphomas | 2075 | 57.20 | 77.26 | 67.51 | 59.81 | 9.83 | 2.62 | 5.07 | 55.81 | 74.34 | 62.90 | 3.53 | 5.99 |
| III CNS tumours | 1356 | 39.01 | 62.50 | 51.81 | 52.29 | 6.64 | 2.44 | 2.11 | 44.25 | 70.02 | 49.40 | 3.61 | 4.22 |
| IV Neuroblastoma | 440 | 50.91 | 74.48 | 66.02 | 54.55 | 10.91 | 2.67 | 2.91 | 53.18 | 75.37 | 64.80 | 2.67 | 3.88 |
| V Retinoblastoma | 172 | 64.16 | 79.39 | 53.66 | 65.32 | 7.51 | 6.11 | 2.44 | 57.56 | 77.10 | 60.98 | 9.16 | 12.20 |
| VI Renal tumours | 738 | 54.88 | 78.22 | 64.39 | 57.99 | 9.89 | 1.32 | 3.03 | 57.32 | 76.40 | 56.82 | 2.15 | 2.27 |
| VII Hepatic tumours | 61 | 52.46 | 74.42 | 50.00 | 49.18 | 3.28 | 0.00 | 5.56 | 44.26 | 62.79 | 44.44 | 0.00 | 5.56 |
| VIII Bone tumours | 588 | 15.65 | 59.48 | 41.13 | 34.18 | 6.63 | 1.51 | 0.00 | 45.92 | 72.84 | 48.39 | 3.66 | 1.61 |
| IX Soft tissue sarcomas | 645 | 47.93 | 72.78 | 60.80 | 53.14 | 8.73 | 1.51 | 0.80 | 50.61 | 73.35 | 56.00 | 4.91 | 4.00 |
| X Germ cell tumours | 386 | 50.26 | 73.31 | 57.33 | 54.14 | 7.51 | 0.96 | 9.33 | 50.52 | 76.53 | 50.67 | 2.25 | 10.67 |
| XI Epithelial neoplasms & melanomas | 130 | 50.77 | 72.34 | 58.33 | 58.46 | 5.38 | 3.19 | 0.00 | 46.15 | 68.09 | 50.00 | 4.26 | 0.00 |
| Other malignant neoplasms<sup>#</sup> | 114 | 63.16 | 69.64 | 68.97 | 66.67 | 14.91 | 7.14 | 1.72 | 55.26 | 75.00 | 60.34 | 5.36 | 3.45 |
| Age at survey (years) |  |  |  |  |  |  |  |  |  |  |  |  |  |
| 18-<25 | 1636 | 56.91 | 70.51 | 62.50 | 56.91 | 10.70 | 3.46 | 2.10 | 52.57 | 70.77 | 57.71 | 4.62 | 4.21 |
| 25-<30 | 2997 | 56.26 | 76.40 | 64.99 | 62.30 | 10.14 | 3.00 | 4.02 | 57.26 | 74.28 | 56.74 | 3.56 | 4.43 |
| 30-<35 | 2463 | 53.76 | 75.05 | 61.33 | 58.02 | 9.38 | 2.39 | 3.93 | 53.02 | 74.44 | 59.21 | 4.17 | 3.32 |
| 35-<40 | 1437 | 46.97 | 70.75 | 63.13 | 44.30 | 6.47 | 1.43 | 4.47 | 52.12 | 74.04 | 62.57 | 2.78 | 6.15 |
| 40-<45 | 833 | 37.45 | 65.05 | 55.07 | 33.05 | 4.20 | 1.57 | 5.80 | 46.58 | 71.07 | 56.52 | 2.49 | 8.70 |
| >=45 | 505 | 26.14 | 60.81 | 52.63 | 56.28 | 4.55 | 2.14 | 7.89 | 42.38 | 72.38 | 57.89 | 5.57 | 10.53 |

Abbreviations: BP: bodily pain, GH: general health, ICCC-3: International Classification of Childhood Cancer, 3rd edition, MH: mental health, n/a: not applicable, PF: physical functioning, RE: role emotional, RP: role physical, SF: social functioning, V1: Version 1, V2: Version 2, VT: vitality, SF-36: Short Form-36.
\* Values calculated separately for V1 and V2, as number of response choices differs between versions.
<sup>#</sup> ICCC-3 main group XII (Other and unspecified malignant neoplasms) and Langerhans cell histiocytosis but not benign and in situ tumours and tumour-like lesions or unclassified survivors.

### Item-internal consistency

In the total cohort and stratified by country, version, gender, and age at survey, all correlations were above the threshold of 0.4, indicating satisfactory consistency between items within a scale (**Table 3**). We found the lowest correlation coefficients among survivors in the Czech Republic for 5 out of 8 scales (PF, RP, BP, SF, RE) and the highest coefficients in Germany (PF, BP, SF) and France (RP, GH, VT, RE). Item-rest correlations were lower in V1 than in V2 for RP (0.67–0.75 vs. 0.80–0.84) and RE (0.65– 0.70 vs. 0.75–0.82). For survivors of some cancers, correlations were below 0.4: for survivors of hepatic tumours and neuroblastoma, correlations within PF were 0.35 for item 3f and 0.38 for item 3j, respectively. For survivors of epithelial neoplasms and melanomas, and other malignant neoplasms, correlations within MH were 0.36 for item 9b (V2), and 0.36 for item 9d (V2).

**Table 3:** Ranges of item-internal consistency correlation coefficients (item-rest correlations) by country, SF-36 version, gender, cancer diagnosis, and age at survey.

|  | n | PF | RP* | BP | GH | VT* | SF | RE* | MH* |
| --- | --- | --- | --- | --- | --- | --- | --- | --- | --- |
| Overall cohort | 9871 | 0.56–0.79 | 0.67–0.84 | 0.87 | 0.54–0.75 | 0.63–0.69 | 0.74 | 0.65–0.82 | 0.59–0.68 |
| **Country** |  |  |  |  |  |  |  |  |  |
| Czech Republic | 1029 | 0.46–0.76 | 0.61–0.72 | 0.83 | 0.56–0.78 | 0.56–0.78 | 0.68 | 0.59–0.70 | 0.60–0.69 |
| France | 385 | 0.61–0.76 | 0.83–0.88 | 0.87 | 0.54–0.86 | 0.54–0.80 | 0.72 | 0.77–0.83 | 0.53–0.73 |
| Germany | 4726 | 0.56–0.81 | 0.65–0.74 | 0.89 | 0.50–0.73 | 0.64–0.66 | 0.77 | 0.64–0.67 | 0.62–0.76 |
| The Netherlands | 2146 | 0.54–0.79 | 0.74–0.79 | 0.85 | 0.57–0.80 | 0.57–0.70 | 0.73 | 0.69–0.77 | 0.54–0.78 |
| Switzerland | 1585 | 0.59–0.79 | 0.79–0.83 | 0.83 | 0.48–0.78 | 0.48–0.74 | 0.71 | 0.75–0.81 | 0.57–0.68 |
| **Version of SF–36** |  |  |  |  |  |  |  |  |  |
| 1 | 7901 | 0.54–0.79 | 0.67–0.75 | 0.87 | 0.54–0.75 | 0.63–0.67 | 0.74 | 0.65–0.70 | 0.59–0.76 |
| 2 | 1970 | 0.61–0.77 | 0.80–0.84 | 0.85 | 0.52–0.76 | 0.64–0.69 | 0.72 | 0.75–0.82 | 0.59–0.68 |
| **Gender** |  |  |  |  |  |  |  |  |  |
| Male | 4725 | 0.60–0.80 | 0.65–0.84 | 0.86 | 0.55–0.74 | 0.60–0.67 | 0.74 | 0.65–0.84 | 0.54–0.75 |
| Female | 5146 | 0.53–0.76 | 0.68–0.86 | 0.87 | 0.53–0.76 | 0.59–0.70 | 0.74 | 0.65–0.81 | 0.60–0.76 |
| **Cancer diagnosis (ICCC-3)** |  |  |  |  |  |  |  |  |  |
| I Leukaemias | 3157 | 0.61–0.77 | 0.66–0.83 | 0.87 | 0.53–0.75 | 0.62–0.67 | 0.72 | 0.61–0.86 | 0.55–0.75 |
| II Lymphomas | 2075 | 0.45–0.75 | 0.64–0.83 | 0.86 | 0.56–0.75 | 0.65–0.75 | 0.75 | 0.65–0.86 | 0.57–0.77 |
| III CNS tumours | 1356 | 0.66–0.82 | 0.67–0.85 | 0.86 | 0.53–0.74 | 0.54–0.64 | 0.74 | 0.68–0.80 | 0.58–0.75 |
| IV Neuroblastoma | 440 | 0.38–0.73 | 0.59–0.89 | 0.84 | 0.53–0.77 | 0.61–0.74 | 0.69 | 0.56–0.86 | 0.57–0.74 |
| V Retinoblastoma | 172 | 0.44–0.80 | 0.56–0.91 | 0.89 | 0.52–0.75 | 0.60–0.77 | 0.80 | 0.73–0.83 | 0.63–0.84 |
| VI Renal tumours | 738 | 0.52–0.74 | 0.64–0.87 | 0.88 | 0.51–0.76 | 0.54–0.69 | 0.76 | 0.66–0.80 | 0.58–0.78 |
| VII Hepatic tumours | 61 | 0.35–0.77 | 0.65–0.95 | 0.84 | 0.52–0.81 | 0.49–0.86 | 0.85 | 0.53–0.86 | 0.63–0.83 |
| VIII Bone tumours | 588 | 0.41–0.79 | 0.72–0.90 | 0.87 | 0.48–0.75 | 0.53–0.80 | 0.75 | 0.71–0.77 | 0.59–0.75 |
| IX Soft tissue sarcomas | 645 | 0.46–0.76 | 0.73–0.88 | 0.86 | 0.56–0.79 | 0.64–0.74 | 0.75 | 0.66–0.83 | 0.50–0.77 |
| X Germ cell tumours | 386 | 0.53–0.83 | 0.71–0.85 | 0.88 | 0.54–0.76 | 0.53–0.75 | 0.74 | 0.66–0.74 | 0.60–0.75 |
| XI Epithelial neoplasms & melanomas | 130 | 0.47–0.85 | 0.73–0.83 | 0.88 | 0.50–0.75 | 0.46–0.75 | 0.72 | 0.71–0.81 | 0.36–0.77 |
| Other malignant neoplasms<sup>#</sup> | 114 | 0.62–0.85 | 0.68–0.94 | 0.87 | 0.71–0.75 | 0.53–0.79 | 0.71 | 0.51–0.88 | 0.36–0.77 |
| **Age at survey (years)** |  |  |  |  |  |  |  |  |  |
| <25 | 1636 | 0.61–0.76 | 0.57–0.83 | 0.81 | 0.53–0.75 | 0.59–0.71 | 0.68 | 0.63–0.81 | 0.50–0.73 |
| 25–30 | 2997 | 0.58–0.78 | 0.62–0.85 | 0.87 | 0.52–0.73 | 0.64–0.70 | 0.75 | 0.60–0.81 | 0.56–0.75 |
| 30–35 | 2463 | 0.53–0.78 | 0.66–0.83 | 0.88 | 0.53–0.73 | 0.65–0.74 | 0.75 | 0.67–0.87 | 0.61–0.78 |
| 35–40 | 1437 | 0.56–0.81 | 0.66–0.86 | 0.87 | 0.53–0.73 | 0.61–0.71 | 0.73 | 0.68–0.83 | 0.57–0.76 |
| 40–45 | 833 | 0.54–0.82 | 0.77–0.94 | 0.9 | 0.53–0.76 | 0.60–0.86 | 0.76 | 0.69–0.93 | 0.56–0.76 |
| >45 | 505 | 0.51–0.79 | 0.78–0.96 | 0.85 | 0.57–0.78 | 0.51–0.85 | 0.75 | 0.71–0.93 | 0.53–0.80 |

Abbreviations: PF: physical functioning, RP: role physical, BP: bodily pain, GH: general health, VT: vitality, SF: social functioning, RE: role emotional, MH: mental health.
ICCC-3: International Classification of Childhood Cancer, 3rd edition, SF-36: Short Form-36.
Note: Rows indicate the lowest and highest coefficients for each scale with $\geq 2$ items (range). The scales BP and SF consist of only 2 items.
\*Values calculated separately for Version 1 and Version 2, as number of response choices differs between versions.
<sup>#</sup>ICCC-3 main group XII (Other and unspecified malignant neoplasms) and Langerhans cell histiocytosis but not benign tumours, in situ tumours, tumour-like lesions or unclassified survivors.

### Item-discriminant validity

Overall, all items correlated better with the items in their hypothesized scale than with the items in unrelated scales (data not shown). However, we found exceptions when we stratified the sample by country or cancer diagnosis. For example, in Switzerland and France—the countries using V2—item 9h (MH) correlated slightly better with VT; in France and The Netherlands, item 9a (VT) correlated slightly better with MH. Among survivors of retinoblastoma, hepatic tumours, epithelial neoplasm and melanoma, and other tumours, few items correlated better with unrelated scales than with their hypothesized scale. In retinoblastoma survivors, item 1 (GH) correlated slightly higher with SF; item 3a (PF) with BP, SF, and GH; and item 9g (VT) with MH. In survivors of hepatic tumours, item 3g (PF) correlated higher with SF; and items 9a, 9e (VT) with MH. In survivors of epithelial neoplasm and melanoma, item 9a (VT) correlated higher with MH. In survivors of other tumours, item 3a (PF) correlated higher with GH; and item 9a (VT) with MH. These differences never exceeded 2 standard errors, so we concluded 100% scaling success for all items in all subgroups.

### Reliability (Cronbach’s alpha)

In the total cohort, Cronbach’s alpha ranged from 0.82–0.93, which was higher than the suggested threshold of 0.7 for scale reliability (**Table 4**) [11]. Reliability differed depending on country, version, gender, age at survey, and diagnostic group. Survivors in the Czech Republic showed slightly lower values for Cronbach’s alpha on most scales, yet higher values for GH and VT than survivors from France and Switzerland who used V2. Cronbach’s alpha was slightly higher on most scales in survivors older than 40 years compared to younger survivors. Cronbach’s alpha was slightly higher on most scales in survivors older than 40 years compared to younger survivors.

**Table 4:** Internal consistency measured with Cronbach’s alpha coefficient by country, SF-36 version, gender, cancer diagnosis, and age at survey.

|  | PF | RP* |  | BP | GH | VT* |  | SF | RE* |  | MH* |  |
| --- | --- | --- | --- | --- | --- | --- | --- | --- | --- | --- | --- | --- |
|  |  | V1 | V2 |  |  | V1 | V2 |  | V1 | V2 | V1 | V2 |
| Overall cohort n=9871 | 0.91 | 0.86 | 0.93 | 0.93 | 0.84 | 0.83 | 0.83 | 0.85 | 0.82 | 0.89 | 0.86 | 0.83 |
| Country |  |  |  |  |  |  |  |  |  |  |  |  |
| Czech Republic | 0.90 | 0.84 | n/a | 0.90 | 0.85 | 0.87 | n/a | 0.80 | 0.79 | n/a | 0.84 | n/a |
| France | 0.90 | n/a | 0.94 | 0.93 | 0.86 | n/a | 0.81 | 0.84 | n/a | 0.89 | n/a | 0.83 |
| Germany | 0.92 | 0.85 | n/a | 0.94 | 0.83 | 0.83 | n/a | 0.87 | 0.80 | n/a | 0.87 | n/a |
| The Netherlands | 0.92 | 0.89 | n/a | 0.92 | 0.86 | 0.82 | n/a | 0.84 | 0.86 | n/a | 0.87 | n/a |
| Switzerland | 0.91 | n/a | 0.92 | 0.91 | 0.82 | n/a | 0.83 | 0.83 | n/a | 0.89 | n/a | 0.82 |
| Version |  |  |  |  |  |  |  |  |  |  |  |  |
| 1 | 0.91 | 0.86 | n/a | 0.93 | 0.84 | 0.83 | n/a | 0.85 | 0.82 | n/a | 0.86 | n/a |
| 2 | 0.91 | n/a | 0.93 | 0.94 | 0.83 | n/a | 0.83 | 0.84 | n/a | 0.89 | n/a | 0.83 |
| Gender |  |  |  |  |  |  |  |  |  |  |  |  |
| Male | 0.92 | 0.86 | 0.92 | 0.92 | 0.83 | 0.83 | 0.82 | 0.85 | 0.81 | 0.90 | 0.86 | 0.79 |
| Female | 0.91 | 0.87 | 0.93 | 0.93 | 0.84 | 0.82 | 0.84 | 0.85 | 0.81 | 0.83 | 0.87 | 0.86 |
| Cancer diagnosis (ICCC-3) |  |  |  |  |  |  |  |  |  |  |  |  |
| I Leukaemias | 0.91 | 0.85 | 0.92 | 0.93 | 0.83 | 0.83 | 0.81 | 0.84 | 0.80 | 0.90 | 0.86 | 0.81 |
| II Lymphomas | 0.89 | 0.85 | 0.91 | 0.92 | 0.84 | 0.84 | 0.86 | 0.85 | 0.82 | 0.90 | 0.86 | 0.82 |
| III CNS tumours | 0.93 | 0.88 | 0.92 | 0.92 | 0.84 | 0.79 | 0.80 | 0.85 | 0.84 | 0.89 | 0.86 | 0.84 |
| IV Neuroblastoma | 0.88 | 0.82 | 0.93 | 0.91 | 0.84 | 0.83 | 0.84 | 0.82 | 0.77 | 0.89 | 0.86 | 0.85 |
| V Retinoblastoma | 0.88 | 0.87 | 0.94 | 0.94 | 0.83 | 0.82 | 0.87 | 0.89 | 0.87 | 0.88 | 0.88 | 0.87 |
| VI Renal tumours | 0.87 | 0.87 | 0.93 | 0.93 | 0.85 | 0.83 | 0.80 | 0.86 | 0.81 | 0.87 | 0.87 | 0.84 |
| VII Hepatic tumours | 0.85 | 0.89 | 0.97 | 0.91 | 0.84 | 0.77 | 0.89 | 0.92 | 0.76 | 0.89 | 0.86 | 0.91 |
| VIII Bone tumours | 0.91 | 0.89 | 0.94 | 0.93 | 0.83 | 0.82 | 0.89 | 0.86 | 0.86 | 0.88 | 0.86 | 0.83 |
| IX Soft tissue sarcomas | 0.89 | 0.88 | 0.94 | 0.92 | 0.85 | 0.85 | 0.82 | 0.85 | 0.82 | 0.89 | 0.88 | 0.83 |
| X Germ cell tumours | 0.92 | 0.88 | 0.90 | 0.93 | 0.83 | 0.79 | 0.87 | 0.85 | 0.84 | 0.85 | 0.86 | 0.87 |
| XI Epithelial neoplasms & melanomas | 0.90 | 0.90 | 0.91 | 0.94 | 0.82 | 0.85 | 0.75 | 0.84 | 0.86 | 0.89 | 0.83 | 0.82 |
| Other malignant neoplasms<sup>#</sup> | 0.92 | 0.88 | 0.95 | 0.93 | 0.86 | 0.88 | 0.80 | 0.83 | 0.72 | 0.91 | 0.84 | 0.79 |
| Age at survey (years) |  |  |  |  |  |  |  |  |  |  |  |  |
| <25 | 0.90 | 0.81 | 0.92 | 0.90 | 0.83 | 0.83 | 0.80 | 0.81 | 0.80 | 0.88 | 0.84 | 0.82 |
| 25–30 | 0.91 | 0.83 | 0.93 | 0.93 | 0.83 | 0.83 | 0.84 | 0.86 | 0.78 | 0.88 | 0.86 | 0.82 |
| 30–35 | 0.91 | 0.86 | 0.92 | 0.93 | 0.84 | 0.83 | 0.86 | 0.86 | 0.82 | 0.91 | 0.87 | 0.87 |
| 35–40 | 0.92 | 0.87 | 0.92 | 0.93 | 0.84 | 0.82 | 0.84 | 0.84 | 0.83 | 0.90 | 0.87 | 0.84 |
| 40–45 | 0.92 | 0.91 | 0.96 | 0.95 | 0.84 | 0.82 | 0.91 | 0.86 | 0.85 | 0.95 | 0.88 | 0.84 |
| >45 | 0.92 | 0.89 | 0.97 | 0.92 | 0.85 | 0.82 | 0.91 | 0.85 | 0.88 | 0.94 | 0.87 | 0.85 |

Abbreviations: PF: physical functioning, RP: role physical, BP: bodily pain, GH: general health, VT: vitality, SF: social functioning, RE: role emotional, MH: mental health.
ICCC-3: International Classification of Childhood Cancer, 3rd edition. n/a: not applicable, SF-36: Short Form-36.
<sup>#</sup>ICCC-3 main group XII (Other and unspecified malignant neoplasms) and Langerhans cell histiocytosis but not benign tumours, in situ tumours, tumour-like lesions or unclassified survivors.

We calculated values separately for versions 1 and 2, since response choices differed between versions.

CCC-3 main group XII (Other and unspecified malignant neoplasms) and Langerhans cell histiocytosis but not benign tumours, in situ tumours, tumour-like lesions or nclassified survivors.

### Scaling assumptions of the summary scores

Of the 8 SF-36 scales, 4 scales represented strong mental health content, and the other 4 represented strong physical health content. In both versions of the SF-36, the scales VT, SF, RE, and MH loaded high on the mental health summary score MCS; and PF, RP, BP, and GH loaded high on the physical health summary score PCS (**Figure 1**). In V2, GH loaded slightly higher on the MCS than in V1, while the PCS loading stayed similar. The MCS and PCS explained 70% (V1) and 71% (V2) of the variance.

**Figure 1:**
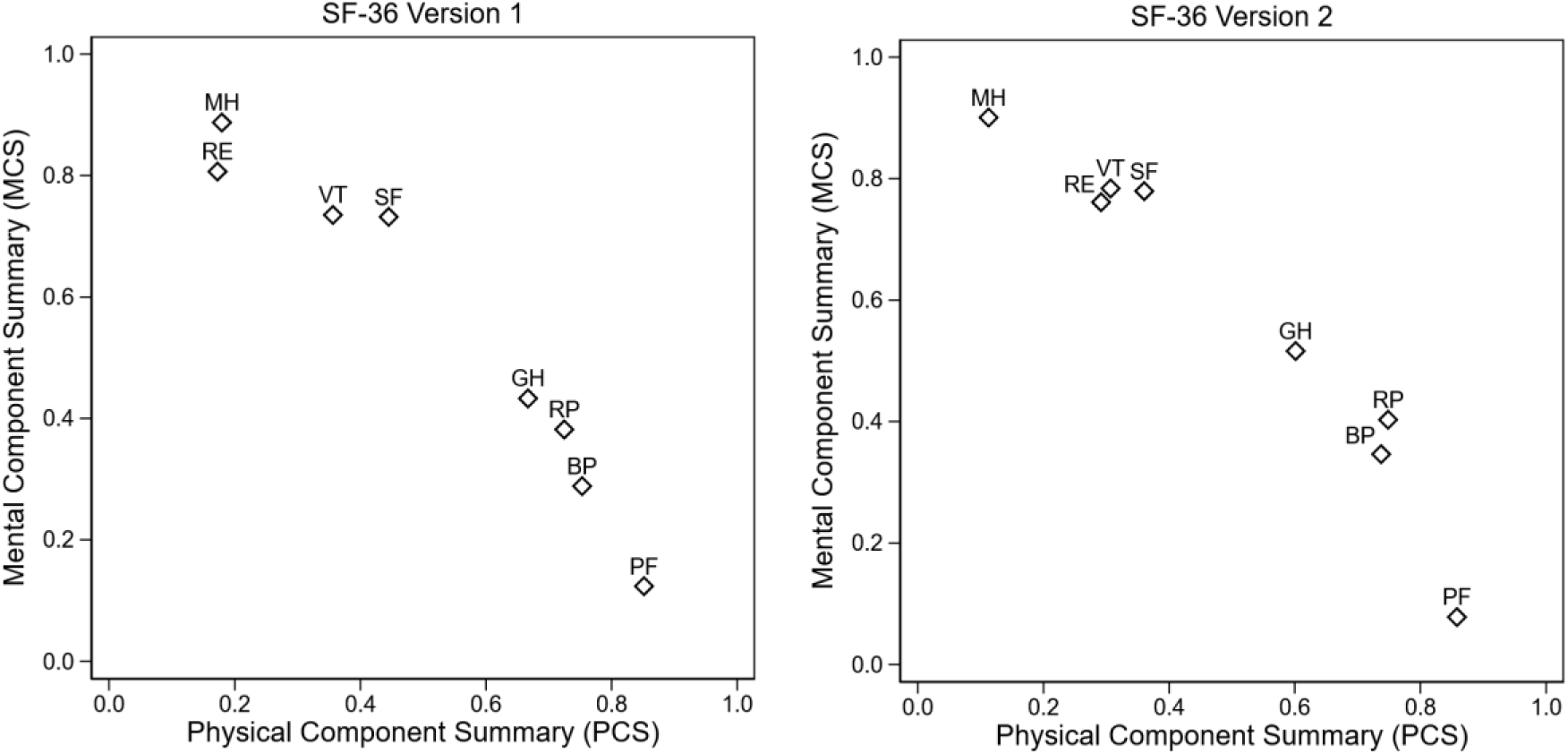
Scaling assumptions of the summary scores; rotated factor loadings for PCS and MCS derived from principal component analyses and orthogonal varimax rotation

## DISCUSSION

In our study of childhood cancer survivors from 5 European countries, we found the SF-36 providing satisfactory measurement properties in the pooled cohort. Both versions of the SF-36 seem appropriate to measure HRQOL among childhood cancer survivors, yet the 2 versions possessed some advantages in specific scales. Differences between countries and versions were generally minor, which justifies combining cohorts from different countries for standard analyses of HRQOL by considering the clustering of participants within countries.

### Data completeness

Data completeness was high for all countries and comparable to the BCCSS [25] or to studies in normal populations [20], although it was lowest among survivors from the Czech Republic. France, Germany, The Netherlands, and Switzerland contacted survivors via postal mail. Since the Czech Republic collected data during clinical follow-up visits, they had the highest percentage of responding survivors among all countries. Generally, face-to-face interviews yield better response rates than postal surveys [33], and in the Czech Republic direct contact between survivors and clinicians during clinical follow-up visits possibly further motivated participation. In contrast, survivors from the Czech Republic showed the largest number of unanswered questions among all countries possibly from time constraints while completing questionnaires in clinics or the high response rate subsequently leading to more missing individual items.

### High ceiling effects

In 5 of 8 scales (PF, RP, BP, SF, RE), high ceiling effects may limit the ability to differentiate among survivors who fared particularly well. We suggest researchers interpret minor HRQOL differences among these participants with caution. RP and RE showed exceptionally high ceiling effects. In a study comparing SF-36 properties across 11 countries, RP and RE ceiling effects were also high (63–83%% and 69–83%, respectively) [20]. Similarly high ceiling effects occurred in the BCCSS [25] and for normal populations in many countries [19; 21-23; 34; 35]. V1 offered 2 response choices for each item, while V2 used a 5-point Likert scale. The latter improved precision for these 2 scales, yet ceiling effects were still high (>15%). Across all scales, men demonstrated higher ceiling effects compared with women. We found ceiling effects decreased with age, which the BCCSS also found [25]. Older age and female gender are well-established risk factors for reporting decreased HRQOL [18; 23; 34]. Again, similar to the BCCSS, survivors of bone tumours showed the lowest ceiling effects in physical scales [25], likely because they are among those with the highest risk for late effects [36]. Floor effects were negligible; thus, SF-36 seems to distinguish well among survivors who score the lowest — except for a reduced reliability in PF — for the most vulnerable groups of survivors of bone tumours or older adult survivors.

### Item-internal consistency, item-discriminant validity and reliability (Cronbach’s alpha)

We observed adequate item-internal consistency, item-discriminant validity, and reliability (Cronbach’s alpha), indicating that the items within a scale measured the concept of its own scale and not the concepts of other scales. However, we noted a few exceptions for item-internal consistency; in the scales PF and MH values were slightly below 0.4 for survivors of hepatic tumours and neuroblastoma (PF), and for survivors of epithelial neoplasms and melanomas (MH), yet with negligible deviations from thresholds and for only a few items of the respective scale. This may be due to low numbers of survivors in these diagnostic subgroups. Our results align with the good consistency and validity reported by the BCCSS [25].

### Tests of scaling assumptions

With principal component analysis, the tests of scaling assumptions indicated that PCS and MCS describe self-assessed physical and mental aspects of HRQOL in European European survivors rather well. Reulen et al. reported a similar explained variance (70%) by the 2 summary scores for the BCCSS [25]. As expected, the scales PF, RP, BP, and GH loaded highest on the PCS, and the scales VT, SF, RE, and MH loaded highest on the MCS. The scale GH loaded slightly higher on the PCS than on the MCS, yet it considerably contributed to both summary scores. This is not surprising, as the developers of the SF-36 intended GH to be sensitive to both physical and mental health, but to correlate higher with the PCS [11]. Compared with V1, GH loaded slightly higher on the MCS in V2. As GH-related items did not change between versions, differences are likely due to different characteristics of survivors between countries, for example, a younger age in survivors from countries using V2 than in countries using V1 rather than a difference related to the two SF-36 versions. We also cannot exclude that differences in GH reflect different cultural perceptions between countries.

### Comparisons between different versions of the SF-36

SF-36 developers [11] and studies from Germany, the United Kingdom, Sweden, and Australia [22; 24; 35; 37] compared both SF-36 versions and found them comparable, yet V2 performed slightly better than the V1 with fewer missing items; decreased ceiling and floor effects; and increased internal consistency (reliability). We observed only less pronounced ceiling effects in V2 than in V1; the other measurement properties were comparable in both versions. Since France and Switzerland were the only countries using V2 of the SF-36, it was not possible to distinguish the effects of version from an effect of country. Switzerland is a country with a high general HRQOL [23], which possibly translates to high ceiling effects. Switzerland had the highest ceiling effects and also contributed most participants using V2. Switzerland showed the highest ceiling effects in scales that are the same in both versions (PF, BP, GH, SF). In changed scales for V2 (RP, RE), ceiling effects in Switzerland were second lowest behind France—indicating lower ceiling effects for V2 compared to V1 of the SF-36.

### Strengths and limitations

Our study exhibits several strengths. Since we assessed measurement properties of the SF-36 (V1 and V2) in a large and diverse cohort of survivors in Europe, it allowed comparing measurement properties between questionnaire versions and countries. A central data centre within PanCareLIFE provided a pooled and harmonized dataset to avoid technical inconsistencies between and within datasets from the 5 countries [26; 38]. In preparation of the study, expert committees of clinicians, biostatisticians, and patient representatives approved all procedures, including study design, data collection, management, analyses of data, and publication of results. Yet, country-specific differences in study design were unavoidable, possibly leading to selection bias [26]. However, such differences also provided opportunities. Although different countries administered different SF-36 versions, assessed HRQOL during different periods (between 2005–2017), and different study questionnaire sections presented the SF-36 questions, we found SF-36 a robust tool to assess HRQOL among childhood cancer survivors.

PanCareLIFE participants answered questionnaires over a span of 12 years [26], which might limit our comparisons of floor and ceiling effects between countries or SF-36 versions because HRQOL or HRQOL perception possibly changed over time. Maruish et al. compared SF-36 raw scores and T-scores scores between 1998 and 2009 US normative samples [11]. HRQOL was slightly higher in 2009 than in 1998—except for a few exceptions on the BP and GH scales—with no clinically meaningful differences between 1998 and 2009 data. Therefore, we believe that our analyses are valid, even when considering the long time span of data collection. In studies investigating changes in HRQOL over time, more sensitive measuring tools might perform better.

## Conclusion

Our results show that both SF-36 versions (V1 and V2) display robust properties to measure HRQOL among childhood cancer survivors in Europe. Nevertheless, we recommend further research to evaluate whether SF-36 proves valid and reliable for intercontinental studies. In our pan-European multicenter study, which combined diverse cohorts, we observed valid and reliable measurement properties across 5 the European countries—a reassurance for researchers planning to use the SF-36 as an HRQOL assessment tool in multicenter studies.

## Data Availability

The data that support the information of this manuscript were accessed on secured servers of the Institute of Social and Preventive Medicine at the University of Bern. Individual-level sensitive data can only be made available for researchers who fulfil the respective legal requirements. All data requests should be communicated to the corresponding author.

## DATA CONTRIBUTIONS

Data for this sub-project were provided by Academisch Medisch Centrum bij de Universiteit van Amsterdam, Netherlands (Prof. Dr. LCM Kremer), Stichting VU-VUMC, Amsterdam, Netherlands (Dr. E van Dulmen-den Broeder, Dr. MH van den Berg), Erasmus Medisch Centrum Rotterdam, Netherlands (Prof. Dr. MM van den Heuvel-Eibrink), Prinses Máxima Centrum, Netherlands (Prof. Dr. MM van den Heuvel-Eibrink, Prof. Dr. LCM Kremer, Dr. E van Dulmen-den Broeder), on behalf of the DCOG LATER Study centres, Universität Bern, Switzerland (Prof. Dr. CE Kuehni), Fakultni nemocnice Brno, Czech Republic (Dr. T Kepak), Centre Hospitalier Universitaire Saint Étienne, France (Dr. C Berger), Fakultni nemocnice v Motole, Prague, Czech Republic (Dr. J Kruseova) and Universitaetsklinikum Bonn, Bonn, Germany (Dr. G Calaminus).

## ETHICS DECLARATION

PanCareLIFE is a multinational collaborative research project that has harmonized and combined data from regional and national cohorts across Europe. Data collection and analysis for each cohort was approved by the responsible ethics committee in each participating country. For Germany this is the Ethics Committee of the Medical Association of Westphalia-Lippe and the Medical faculty of the Westphalian Wilhelms University (2012-530-f-S), for Switzerland the Cantonal Ethics Committee of the Canton of Bern (KEK-BE: 166/2014; 2021-01462), for the Czech Republic the Ethics Committee for Multi-Centric Clinical Trials of the University Hospital Motol (EK-1723/13) and the Multi-Centric Ethics Committee of the University Hospital Brno (approval date: 2014/10/22), for France the Personal Protection Committee South East 1 (CPP: 2015-23) and for the Netherlands the Medical Ethical Review Committee of the Academic Medical Center, University of Amsterdam (2010_332#B2015108).

## ABBREVIATIONS

BCCSS: British Childhood Cancer Survivor Study
BP: Bodily pain
CCSS: Childhood Cancer Survivor Study
CNS: Central Nervous System
CHU-SE: Centre Hospitalier Universitaire de Saint-Étienne, St. Étienne, France
DCOG LATER: Dutch Childhood Oncology Group Survivor study
FNM: Motol Teaching Hospital, Prague, Czech Republic
GCCR: German Childhood Cancer Registry
GH: General Health
HRQOL: Health-related Quality of Life
ICCC-3: International Classification of Childhood Cancer, 3rd edition
MCS: Mental Component Summary
MH: Mental health
n/a: not applicable
PCS: Physical Component Summary
PF: Physical functioning
RE: Role-limitations due to emotional problems
RP: Role-limitations due to physical problems
SCCR: Swiss Childhood Cancer Registry
SCCSS: Swiss Childhood Cancer Survivor Study
F: Social functioning
SF-36: Short-Form 36
UHB: University Hospital Brno, Czech Republic
UKB: Universitätsklinikum Bonn, Germany
UKM: Universitätsklinikum Münster, Germany
UNIBE: University of Bern
UK: United Kingdom
US: United States
VIVE: First Basic Survey on Life Situation, State of Health, and Quality of Life of Childhood Cancer Survivors in Germany
VT: Vitality

## ACKNOWLEDGEMENTS

We gratefully acknowledge the participation of survivors of childhood and adolescent cancer and their families in this research.

PanCareLIFE (Grant no. 602030) is a collaborative project in the 7th Framework Program of the European Union. Project partners are Universitätsmedizin der Johannes Gutenberg-Universität Mainz, Germany (PD Dr P Kaatsch, Dr D Grabow); Boyne Research Institute, Drogheda, Ireland (Dr J Byrne, Ms H Campbell); Pintail Ltd., Dublin, Ireland (Mr C Clissmann, Dr K O’Brien); Academisch Medisch Centrum bij de Universiteit van Amsterdam, the Netherlands (Prof Dr LCM Kremer); Universität zu Lübeck, Germany (Prof T Langer); Stichting VU-VUMC, Amsterdam, the Netherlands (Dr E van Dulmen-den Broeder, Dr MH van den Berg); Erasmus Universitair Medisch Centrum, Rotterdam, the Netherlands; and Princess Maxima Center for Pediatric Oncology, Utrecht, the Netherlands (Prof Dr MM van den Heuvel-Eibrink); Charité-Universitätsmedizin Berlin, Germany (Prof Dr A Borgmann-Staudt); Westfälische Wilhelms-Universität Münster, Germany (Prof Dr A am Zehnhoff-Dinnesen); Universität Bern, Switzerland (Prof Dr CE Kuehni); Istituto Giannina Gaslini, Genoa, Italy (Dr R Haupt); Fakultni nemocnice Brno, Czech Republic (Dr T Kepak); Centre Hospitalier Universitaire Saint-Étienne, Saint-Étienne, France (Dr C Berger); Kraeftens Bekaempelse, Copenhagen, Denmark (Dr JF Winther); Fakultni nemocnice v Motole, Prague, the Czech Republic (Dr J Kruseova); Universitaetsklinikum Bonn, Bonn, Germany (Dr G Calaminus, K Baust); and University Hospital Essen, Essen, Germany (Prof U Dirksen). The EU Commission takes no responsibility for any use made of the information set out.

In the Czech Republic, at the University Hospital of Brno we thank the team of collaborators from the Children’s Hospital in Brno, the Masaryk University in Brno and Brno survivors’ association Together Towards a Smile: H. Hrstková, V. Bajčiová, D. Hošnová, Z. Kuttnerová, M. Blanářová, R. Mazúr, I. Mikulec, E. Bučková, L. Červinková, E. Bařinová, I. Krupková, E. Novotná, P. Chloupková, Z. Wimmerová, L. Štrublová and numerous other external collaborators and supporters. At the University Hospital of Prague LTFU care registry, team of the Prague Childhood Cancer Survivor Study: Keslová P, Ganevová M, Reichlová V, Bašeová J, Nováková L, Douchová M, Čepelová M.

In France, the team of the long-term follow-up study (SALTO): Faure-Conter C. (Lyon), Corradini N. (Lyon), Plantaz D. (Grenoble), Tarral E. (Grenoble), Durieu I. (Lyon), Guichard I. (Saint-Etienne), Odier F. (Saint-Etienne), Gauthier N. (Lyon), Métral P. (Lyon), Mercier M. (Lyon), Billet S. (Grenoble), Celette C. (Grenoble), Schiff I. (Grenoble), Loubier A. (Saint-Étienne).

In Germany, we thank the VIVE group: G. Calaminus (Bonn) U Creutzig (Hannover), P Kaatsch (Mainz), T Langer (Lübeck), K. Baust (Bonn), J. Dobke (Berlin) I. Jung (Mainz), I. Kerenyi (Mainz), C. Spix (Mainz), C. Teske (Bonn), M. Zimmermann (Hannover).

In The Netherlands, we thank the DCOG LATER consortium: L.C.M. Kremer, E. van Dulmen-den Broeder, M.M. van den Heuvel-Eibrink, W.J.E. Tissing, J. Loonen, D. Bresters, B. Versluijs, M.A. Grootenhuis, M. van der Heiden-van der Loo, F. van Leeuwen, S.J.C. Neggers, H.J.H. van der Pal, C.M. Ronckers, A.C.H. de Vries, G.O.R Janssens, J. den Hartogh. H.M. van Santen, M.A. Veening, M. Louwerens, S.M.F. Pluijm.

In Switzerland, we thank the Swiss Childhood Cancer Survivor Study team, the Swiss Paediatric Oncology Group data managers, and the Swiss Childhood Cancer Registry study team. We also thank Marcel Zwahlen for statistical advice. We thank Kristin Marie Bivens for her editorial guidance and suggestions.

