## Supplemental Table 1 for "Implementation of the Short-Form 36 (SF-36) in childhood cancer survivors: An analysis on measurement properties across 5 European countries"

**Supplemental Table 1: Sociodemographic and medical characteristics of survivors for the overall cohort and by SF-36 version**

| | V1 and V2 combined N=9871 | | V1 N=7901 | | V2 N=1970 | |
|---|---|---|---|---|---|---|
| | n | % | n | % | n | % |
| ***Sex*** | | | | | | |
| Male | 4725 | 47.9 | 3734 | 47.3 | 991 | 50.3 |
| Female | 5146 | 52.1 | 4167 | 52.7 | 979 | 49.7 |
| ***Age at survey (years)*** | | | | | | |
| 18–<25 | 1636 | 16.6 | 780 | 9.9 | 856 | 43.5 |
| 25–<30 | 2997 | 30.4 | 2500 | 31.6 | 497 | 25.2 |
| 30–<35 | 2463 | 25.0 | 2132 | 27.0 | 331 | 16.8 |
| 35–<40 | 1437 | 14.6 | 1258 | 15.9 | 179 | 9.1 |
| 40–<45 | 833 | 8.4 | 764 | 9.7 | 69 | 3.5 |
| 45–69 | 505 | 5.1 | 467 | 5.9 | 38 | 1.9 |
| ***Age at cancer diagnosis (years)*** | | | | | | |
| 0–<5 | 3201 | 32.4 | 2604 | 33.0 | 597 | 30.3 |
| 5–<10 | 2572 | 26.1 | 2123 | 26.9 | 449 | 22.8 |
| 10–<15 | 3448 | 34.9 | 2793 | 35.3 | 655 | 33.2 |
| 15–18 | 650 | 6.6 | 381 | 4.8 | 269 | 13.7 |
| ***Period of cancer diagnosis*** | | | | | | |
| 1963–<1985 | 2086 | 21.1 | 1746 | 22.1 | 340 | 17.3 |
| 1985–<1995 | 4680 | 47.4 | 3698 | 46.8 | 982 | 49.8 |
| 1995–<2005 | 2792 | 28.3 | 2265 | 28.7 | 527 | 26.8 |
| 2005–2010 | 313 | 3.2 | 192 | 2.4 | 121 | 6.1 |
| ***Time since cancer diagnosis (years)*** | | | | | | |
| 5–<10 | 455 | 4.6 | 164 | 2.1 | 291 | 14.8 |
| 10–<15 | 835 | 8.5 | 550 | 7.0 | 285 | 14.5 |
| 15–<20 | 2179 | 22.1 | 1648 | 20.9 | 531 | 27.0 |
| 20–<25 | 2503 | 25.4 | 1962 | 24.8 | 541 | 27.5 |
| 25–<30 | 2023 | 20.5 | 1802 | 22.8 | 221 | 11.2 |
| 30–<35 | 1302 | 13.2 | 1218 | 15.4 | 84 | 4.3 |
| 35–54 | 574 | 5.8 | 557 | 7.0 | 17 | 0.9 |
| ***Cancer diagnosis (ICCC-3)*** | | | | | | |
| I Leukaemias | 3157 | 32.0 | 2665 | 33.7 | 492 | 25.0 |
| II Lymphomas | 2075 | 21.0 | 1641 | 20.8 | 434 | 22.0 |
| III CNS tumours | 1356 | 13.7 | 1024 | 13.0 | 332 | 16.9 |
| IV Neuroblastoma | 440 | 4.5 | 337 | 4.3 | 103 | 5.2 |
| V Retinoblastoma | 172 | 1.7 | 131 | 1.7 | 41 | 2.1 |
| VI Renal tumours | 738 | 7.5 | 606 | 7.7 | 132 | 6.7 |
| VII Hepatic tumours | 61 | 0.6 | 43 | 0.5 | 18 | 0.9 |
| VIII Bone tumours | 588 | 6.0 | 464 | 5.9 | 124 | 6.3 |
| IX Soft tissue sarcomas | 654 | 6.6 | 529 | 6.7 | 125 | 6.3 |
| X Germ cell tumours | 386 | 3.9 | 311 | 3.9 | 75 | 3.8 |
| XI Epithelial neoplasms & melanomas | 130 | 1.3 | 94 | 1.2 | 36 | 1.8 |
| Other malignant neoplasms [#] | 114 | 1.2 | 56 | 0.7 | 58 | 2.9 |
| ***Subsequent tumour*** | | | | | | |
| Yes | 646 | 6.5 | 587 | 7.4 | 59 | 3.0 |
| No | 9225 | 93.5 | 7314 | 92.6 | 1911 | 97.0 |
| ***Treatment*** | | | | | | |
| Surgery only | 642 | 6.5 | 360 | 4.6 | 282 | 14.3 |
| Radiotherapy only | 49 | 0.5 | 45 | 0.6 | 4 | 0.2 |
| Chemotherapy only | 2086 | 21.1 | 1697 | 21.5 | 389 | 19.7 |
| Surgery and radiotherapy | 285 | 2.9 | 195 | 2.5 | 90 | 4.6 |
| Surgery and chemotherapy | 1737 | 17.6 | 1260 | 15.9 | 477 | 24.2 |
| Radiotherapy and chemotherapy | 1975 | 20.0 | 1800 | 22.8 | 175 | 8.9 |
| Radiotherapy, chemotherapy, and surgery | 1847 | 18.7 | 1401 | 17.7 | 446 | 22.6 |
| No surgery, chemotherapy, or radiotherapy | 46 | 0.5 | 27 | 0.3 | 19 | 1.0 |
| Complete treatment information not available | 1204 | 12.2 | 1116 | 14.1 | 88 | 4.5 |
| ***Haematopoietic stem cell transplantation*** | | | | | | |
| Unknown | 307 | 3.1 | 256 | 3.2 | 51 | 2.6 |
| Yes | 307 | 3.1 | 217 | 2.7 | 90 | 4.6 |
| No | 9257 | 93.8 | 7428 | 94.0 | 1829 | 92.8 |

Abbreviations: SF-36: Short Form-36, V1: Version 1, V2: Version 2, ICCC-3: International Classification of Childhood Cancer, 3rd edition,#: ICCC-3 main group XII (Other and unspecified malignant neoplasms) and Langerhans cell histiocytosis but not benign and in situ tumours and tumour-like lesions or unclassified survivors.
