## Supplemental Table 2 for "Implementation of the Short-Form 36 (SF-36) in childhood cancer survivors: An analysis on measurement properties across 5 European countries"

**Supplemental Table 2: Floor effects: Proportions of survivors with the lowest possible score by country, SF-36 version, gender, cancer diagnosis and age at study.**

| | PF | RP* | | BP | GH | VT* | | SF | RE* | | MH* | |
|---|---|---|---|---|---|---|---|---|---|---|---|---|
| | | V1 | V2 | | | V1 | V2 | | V1 | V2 | V1 | V2 |
| **Overall cohort n=9871** | **0.20** | **7.95** | **0.86** | **0.48** | **0.16** | **0.00** | **0.15** | **0.50** | **8.94** | **0.76** | **0.04** | **0.00** |
| ***Country*** | | | | | | | | | | | | |
| Czech Republic | 0.29 | 6.90 | n/a | 0.29 | 0.19 | 0 | n/a | 0.19 | 9.23 | n/a | 0 | n/a |
| France | 0 | n/a | 1.56 | 0.78 | 0.52 | n/a | 0.52 | 0.78 | n/a | 2.08 | n/a | 0 |
| Germany | 0.17 | 6.16 | n/a | 0.63 | 0.11 | 0 | n/a | 0.47 | 7.77 | n/a | 0.04 | n/a |
| The Netherlands | 0.19 | 12.40 | n/a | 0.37 | 0.28 | 0 | n/a | 0.79 | 11.37 | n/a | 0.05 | n/a |
| Switzerland | 0.32 | n/a | 0.69 | 0.19 | 0.06 | n/a | 0.06 | 0.32 | n/a | 0.44 | n/a | 0 |
| ***Version*** | | | | | | | | | | | | |
| 1 | 0.19 | 7.95 | n/a | 0.52 | 0.16 | 0 | n/a | 0.52 | 8.94 | n/a | 0.04 | n/a |
| 2 | 0.25 | n/a | 0.86 | 0.30 | 0.15 | n/a | 0.15 | 0.41 | n/a | 0.76 | n/a | 0 |
| ***Gender*** | | | | | | | | | | | | |
| Male | 0.17 | 6.00 | 0.61 | 0.32 | 0.17 | 0 | 0.20 | 0.51 | 7.53 | 0.61 | 0.08 | 0 |
| Female | 0.23 | 9.70 | 1.12 | 0.62 | 0.16 | 0 | 0.10 | 0.49 | 10.20 | 0.92 | 0.04 | 0 |
| ***Cancer diagnosis (ICCC-3)*** | | | | | | | | | | | | |
| I Leukaemias | 0.10 | 6.34 | 0.20 | 0.41 | 0.16 | 0 | 0 | 0.38 | 8.14 | 0.20 | 0.04 | 0 |
| II Lymphomas | 0.10 | 6.34 | 0.69 | 0.48 | 0.19 | 0 | 0.23 | 0.29 | 8.53 | 0.69 | 0 | 0 |
| III CNS tumours | 0.81 | 12.40 | 1.20 | 0.74 | 0.15 | 0 | 0 | 1.40 | 11.82 | 1.51 | 0.20 | 0 |
| IV Neuroblastoma | 0.23 | 5.04 | 0.97 | 0.23 | 0.23 | 0 | 0.97 | 0.23 | 6.23 | 0 | 0 | 0 |
| V Retinoblastoma | 0 | 5.34 | 4.88 | 0.58 | 0 | 0 | 0 | 1.16 | 9.92 | 0 | 0 | 0 |
| VI Renal tumours | 0 | 5.94 | 0 | 0.27 | 0.27 | 0 | 0 | 0 | 7.43 | 0 | 0 | 0 |
| VII Hepatic tumours | 0 | 9.30 | 5.56 | 0 | 0 | 0 | 0 | 3.28 | 9.30 | 5.56 | 0 | 0 |
| VIII Bone tumours | 0.17 | 15.95 | 2.42 | 0.51 | 0.17 | 0 | 0.81 | 0.68 | 11.21 | 1.61 | 0 | 0 |
| IX Soft tissue sarcomas | 0 | 8.88 | 0.80 | 0.46 | 0.15 | 0 | 0 | 0.15 | 9.07 | 2.40 | 0 | 0 |
| X Germ cell tumours | 0.26 | 8.68 | 0 | 1.04 | 0 | 0 | 0 | 0.52 | 9.00 | 0 | 0 | 0 |
| XI Epithelial neoplasms & melanomas | 0 | 10.64 | 0 | 0 | 0 | 0 | 0 | 0 | 14.89 | 0 | 0 | 0 |
| Other malignant neoplasms[#] | 0.88 | 10.71 | 1.72 | 0 | 0 | 0 | 0 | 0 | 5.36 | 0 | 0 | 0 |
| ***Age at assessment (years)*** | | | | | | | | | | | | |
| <25 | 0.31 | 5.26 | 0.70 | 0.31 | 0.12 | 0 | 0.12 | 0.37 | 9.23 | 0.47 | 0 | 0 |
| 25–30 | 0.07 | 5.20 | 1.01 | 0.23 | 0.13 | 0 | 0 | 0.43 | 7.08 | 0.80 | 0 | 0 |
| 30–35 | 0.16 | 6.66 | 0.30 | 0.73 | 0.20 | 0 | 0.60 | 0.37 | 8.86 | 0.91 | 0 | 0 |
| 35–40 | 0.21 | 8.66 | 1.12 | 0.49 | 0.14 | 0 | 0 | 0.49 | 9.22 | 1.12 | 0.08 | 0 |
| 40–45 | 0.36 | 15.45 | 2.90 | 0.84 | 0.12 | 0 | 0 | 1.08 | 11.91 | 2.90 | 0.26 | 0 |
| >45 | 0.59 | 18.84 | 2.63 | 0.59 | 0.40 | 0 | 0 | 0.99 | 13.06 | 0 | 0 | 0 |

Abbreviations: BP: bodily pain, GH: general health, ICCC-3: International Classification of Childhood Cancer, 3rd edition, MH: mental health, n/a: not applicable, PF: physical functioning, RE: role emotional, RP: role physical, SF: social functioning, V1: Version 1, V2: Version 2, VT: vitality.
* Values calculated separately for V1 and V2, as number of response choices differs between versions.

[#] ICCC-3 main group XII (Other and unspecified malignant neoplasms) and Langerhans cell histiocytosis but not benign and in situ tumours and tumour-like lesions or unclassified survivors.
